# Evaluating Explanations from AI Algorithms for Clinical Decision-Making: A Social Science-based Approach

**DOI:** 10.1101/2024.02.26.24303365

**Authors:** Suparna Ghanvatkar, Vaibhav Rajan

**Affiliations:** Department of Information Systems and Analytics, National University of Singapore, Singapore (,)

**Keywords:** artificial intelligence, explainability, inter-pretability, clinical decision support, explanation evaluation

## Abstract

Explainable Artificial Intelligence (XAI) techniques generate explanations for predictions from AI models. These explanations can be evaluated for (i) faithfulness to the prediction, i.e., its correctness about the reasons for prediction, and (ii) usefulness to the user. While there are metrics to evaluate faithfulness, to our knowledge, there are no automated metrics to evaluate the usefulness of explanations in the clinical context. Our objective is to develop a new metric to evaluate usefulness of AI explanations to clinicians. Usefulness evaluation needs to consider both (a) how humans generally process explanations and (b) clinicians’ specific requirements from explanations presented by clinical decision support systems (CDSS). Our new scoring method can evaluate the usefulness of explanations generated by any XAI method that provides importance values for the input features of the prediction model. Our method draws on theories from social science to gauge usefulness, and uses literature-derived biomedical knowledge graphs to quantify support for the explanations from clinical literature. We evaluate our method in a case study on predicting onset of sepsis in intensive care units. Our analysis shows that the scores obtained using our method corroborate with independent evidence from clinical literature and have the required qualities expected from such a metric. Thus, our method can be used to evaluate and select useful explanations from a diverse set of XAI techniques in clinical contexts, making it a fundamental tool for future research in the design of AI-driven CDSS.

## I. Introduction

Artificial Intelligence (AI) models, such as deep neural networks, are increasingly being used in healthcare to support data-driven decision-making [1]. However, the black-box nature of these models often leads to ethical concerns and lower user acceptance, resulting in poor adoption of AI-driven Clinical Decision Support Systems (CDSS) [1], [2]. To address these concerns and ensure adherence to data protection regulations, such as the European General Data Protection Regulation (GDPR), explanations are required for model predictions [3]. Explainable AI (XAI) algorithms are being actively developed and used to fulfill this need [4]–[10]. The explanations from many such XAI algorithms are in the form of *importance values* for each input feature. Feature importance is a measure of how much each input feature contributes to the output of a prediction model [11]. It can help users understand which features are most relevant for the model’s predictions. E.g., consider a model that takes a brain MRI as input and predicts presence/absence of a disease – an explanation will highlight the 3D voxels in the input MRI that are relevant for the prediction. Feature importance can be computed using various XAI methods such as SHAP [11], LIME [12], LRP [13] etc.

However, for developing AI *predictive systems* (i.e., systems that generate *both* predictions and explanations) for clinical tasks, choosing an XAI algorithm is challenging. This difficulty stems from a large number of choices and the lack of a rigorous metric to evaluate explanations generated by them [4], [14]–[16]. E.g., a recent review on XAI techniques for clinical tasks lists *174* different methods used [15]. These include *interpretable models* that inherently produce explainable predictions (e.g., decision trees), as well as *post-hoc* techniques that produce an explanation after the prediction is generated (e.g., SHAP [11] and LIME [12]). Explanation evaluation can be categorized on two aspects: (a) *faithfulness* of the explanation to the prediction and (b) *usefulness* of the explanation to the user [17]. Faithfulness is the ability of the explanation to correctly represent the reason for the prediction, i.e., whether the important features indicated by the explanation are indeed responsible for the prediction. Usefulness is the ability of the explanation to aid subsequent decision-making of the user in practical settings, implying it is dependent on the use case and user.

We identify four gaps in the literature on the evaluation of explanations from XAI algorithms. First, extant evaluation metrics have focussed mainly on *faithfulness*, e.g., the stability metric evaluates whether similar examples have consistently similar explanations [18]. Second, metrics that evaluate usefulness of explanations may not easily apply to diverse data types and explanations within the same use-case, e.g., [19] provides three metrics for rule-style explanation over text: explanation size, number of cognitive chunks in the explanation, and repeated terms in the explanation, but the latter two may not extend beyond text data based explanations. Third, evaluation of usefulness involves user feedback (e.g., [20], [21]), which is difficult to automate and scale to a large number of users. Fourth, existing metrics have been developed with little involvement and consideration of end-users of the explanations [22]. As the usefulness of explanation depends on the use-case and the user, it must account for end-users’ expectations from the explanation, e.g., clinicians expect explanations in terms of clinical concepts that enables them to effectively utilize the predictions in time-critical decision-making [23]–[26]. We refer to ‘clinical concepts’ as the concepts defined in the Unified Medical Language System (UMLS) [27]. Each clinical concept is represented by a group of terms sharing synonymous meanings; terms being sourced from various standardized medical vocabularies such as SNOMED-CT and ICD. To our knowledge, there are no data-type independent metrics that can evaluate the usefulness of explanations from XAI algorithms in a quantitative, automated, and scalable manner in clinical applications [16], [28].

In this paper, we develop a new scoring method to evaluate the usefulness of explanations (represented as feature-importance values) generated by an AI predictive system, for a clinical task, which addresses the aforementioned gaps. To develop our approach, we turn to theories of explanation from the social sciences that explain how people generate and evaluate explanations in general. This provides a strong theoretical foundation to specify requirements for a useful explanation, which are evaluated through our method. To account for clinicians’ expectations from the explanation, we evaluate them in terms of clinical concepts and also check for agreement with clinical literature through the use of literature-derived biomedical knowledge graphs (KGs). These KGs are large, heterogeneous graphs with multiple node types representing clinical concepts (e.g., diseases, drugs) from standardized vocabularies and multiple edge types (e.g., ‘treats’, ‘predisposes’) representing associations between pairs of clinical concepts. The knowledge in the clinical literature is encapsulated in KG using triples of the form (source node, edge type, target node), such as (High Creatinine, Predisposes, Pulmonary Embolism). The key idea of using KGs is to give higher scores to those explanations that have support from clinical literature. This obviates the need for manual user feedback, making our method automated and scalable. Further, as our evaluation is in terms of clinical concepts, it is independent of the input datatype and can be used for any explanation that can be converted to feature importance values. A preliminary version of this work was presented in [29].

In this paper, we first review the literature on existing evaluation techniques for explanations and the literature of explainability in clinical contexts in Section II. We then describe our methodology to evaluate the explanations generated by predictive systems using the theories of social science and contextualizing them to the clinical setting in Section III.A. We demonstrate the utility of our method through a case study of an AI predictive system that predicts onset of sepsis in the next 6 hours in Intensive Care Units (ICU). The experimental setup is described in Section III.B, while Section IV describes the results of our experiments. Our analysis shows that the scores obtained using our approach corroborates with independent evidence from clinical literature, and satisfies clinicians’ requirements. Lastly, in Section V, we discuss the implications and scope for future work. Notably, we conclude that along with metrics for faithfulness of explanations, our usefulness metric can be used to select a suitable XAI algorithm for a predictive system. A predictive system with a higher score on our metric is expected to provide literature-supported explanations that are useful for clinicians in their decision-making. Thus our metric can potentially lead to improved design and higher adoption of AI-driven CDSS.

## II. Related Work

### A. Evaluating XAI Techniques

Evaluation of explanations can be categorized based on two aspects, viz., *faithfulness* of the explanation to the prediction and *usefulness* of the explanation to the user [17]. Many strategies for evaluating faithfulness have been proposed, such as those based on similarity [30], stability [18], fidelity [31], and modality importance correlation [32]. These evaluation strategies have been reviewed by [20], [28], [33]. These metrics are useful to ensure the explanations represent the predictions correctly, but may not be indicative of usefulness in real-world applications [17].

Many studies evaluate the usefulness of explanations by conducting user-studies [20], [34]–[36] and rely on an existing human-annotated dataset for checking the usefulness [37]. However, such evaluations do not scale well due to their dependence on human observers for evaluation. Reference [19] evaluate the usefulness of explanations for text data in terms of their explanation size and the cognitive load (through the number of cognitive chunks and number of repeated terms). Similarly, [38] evaluate the explanations generated on the biomedical text based on its faithfulness to the prediction and the usefulness by focusing on the number of explanation units (features), which would be representative of the cognitive load. These metrics are specific to text or rule-based explanations, which may not necessarily translate to other data types. Further, all these criteria generally use the input features (as used by the XAI method), which may not be the most appropriate granularity for evaluating an explanation. For example, if the task is to predict a disease using radiology images of the brain, then the features may be in terms of 3D voxels. However, the decision-making process of clinicians, the users of the predictive algorithm and its explanations, may be at the level of brain regions (not specific locations in the brain represented by the voxels). To summarize, extant approaches lack strategies to evaluate the usefulness of explanations in an objective and scalable manner.

### B. Explainability in Clinical Contexts

Usefulness of explanations is dependent on the use-case and the users’ expectations. Different user groups can have different expectations of explanations, depending on the use-case and the subsequent usage of the explanation [39]. For the clinical setting, the end-users (i.e., clinicians) have specific requirements from the systems: they expect systems to demonstrate an understanding of the clinical concepts relevant for subsequent decision-making [23]–[26]. Thus, explanations from predictive systems would be considered useful when they are in terms of clinical concepts and are aligned with their understanding of clinical literature and practice.

The extant literature on XAI in clinical and biomedical context, often do not perform any quantitative evaluation of the different explanations generated (e.g. [6], [8], [9]). Previous reviews on XAI in clinical context, (e.g., [15], [16]), also highlight that studies often do not perform any quantitative evaluation, and the ones evaluating usefulness did so through experiments, interviews, or surveys.

As discussed earlier, most extant literature on evaluating explanations focuses on explanations being faithful to the prediction model. However, within clinical settings, where time-sensitive decisions carry high stakes for clinicians, the criterion of usefulness becomes paramount [24]. Thus, there is a critical need for evaluation methods that can evaluate the usefulness of explanations in an automated manner at the explanation granularity of clinical concepts.

Table I compares the current literature along these dimensions. We note that only [30], [32] focus on the clinical context. Reference [37] defines ‘plausibility’ as the assessment of explanation quality by humans, akin in principle to our notion of usefulness. However, it is designed for images and requires human annotation effort. This metric has also been subsequently used by [32], who not only assess plausibility using annotated data on radiology images, but also introduce a metric for assessing explanation faithfulness to the model. However, based on Table I, a notable gap emerges: there is a dearth of automated evaluation strategies specifically designed for assessing the usefulness of explanations in clinical contexts at the granular level of clinical concepts.

**TABLE I.**
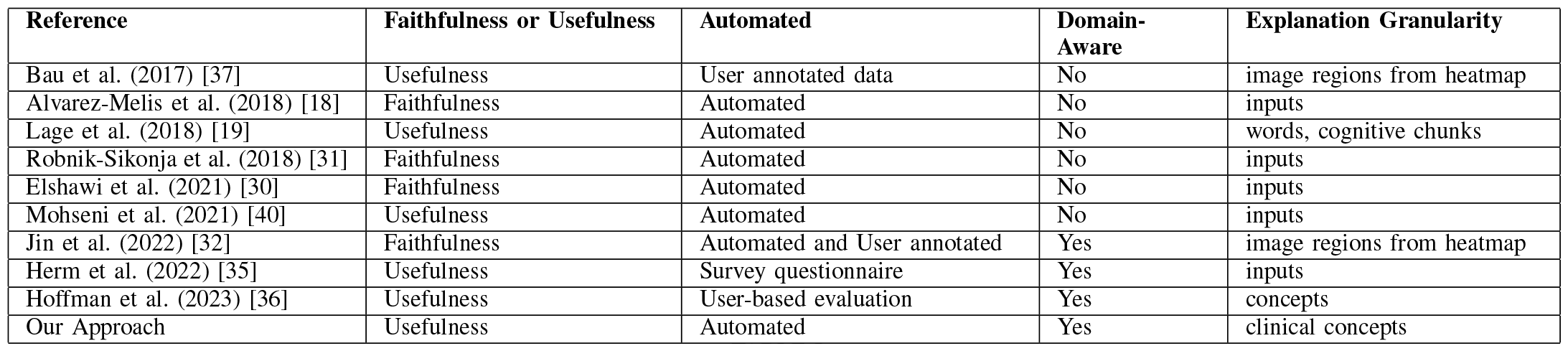
Comparison of our work with recent literature.

| Reference | Faithfulness or Usefulness | Automated | Domain-Aware | Explanation Granularity |
| --- | --- | --- | --- | --- |
| Bau et al. (2017) [37] | Usefulness | User annotated data | No | image regions from heatmap |
| Alvarez-Melis et al. (2018) [18] | Faithfulness | Automated | No | inputs |
| Lage et al. (2018) [19] | Usefulness | Automated | No | words, cognitive chunks |
| Robnik-Sikonja et al. (2018) [31] | Faithfulness | Automated | No | inputs |
| Elshawi et al. (2021) [30] | Faithfulness | Automated | No | inputs |
| Mohseni et al. (2021) [40] | Usefulness | Automated | No | inputs |
| Jin et al. (2022) [32] | Faithfulness | Automated and User annotated | Yes | image regions from heatmap |
| Herm et al. (2022) [35] | Usefulness | Survey questionnaire | Yes | inputs |
| Hoffman et al. (2023) [36] | Usefulness | User-based evaluation | Yes | concepts |
| Our Approach | Usefulness | Automated | Yes | clinical concepts |

## III. Materials and Methods

### A. Our Method

Explanation is a justification or reason for a belief or action or decision; it has been studied extensively in the social sciences. Miller [39] consolidates the frameworks of explanations from social sciences, based on how people define, generate, select, evaluate, and present explanations, to provide four requirements of a *good* explanation: an explanation must (1) be contrastive in nature, (2) have select number of concepts, (3) generate causal understanding, and (4) consider the user.

The first requirement is that explanation must be contrastive,i.e., answer the question ‘why P rather than Q?’; we refer to P as *fact* and Q as *foil*. Selective explanations that provide a contrastive explanation for a particular prediction help develop a causal understanding. We use biomedical knowledge graphs (KG) to quantify the association between the fact/foil and clinical concepts used in or derived from the explanation. These KG-based estimates are used to score a given explanation, as described below. We present these desiderata along with the subsequent design choices leading to the definitions of 5 context-specific variables used in our method in Table II. Fig 1 visualizes the entire evaluation methodology.

**TABLE II.** Design Choice from Desiderata for Usefulness Metric.

| Desiderata | Details | Design Choice | Variables | Choice of Variables |
| --- | --- | --- | --- | --- |
| Explanations are **contrastive** | Explanations do not seek the answer of ‘Why P?’ but rather ‘Why P rather than Q?’, though Q may often be implicit in the context. Following Lipton [41], we refer to P as the *fact* and Q as the *foil*. | Identify the *foils* for the given prediction task, where the *fact* corresponds to the prediction task itself. | i) *fact* statement and ii) *foil* statement | There may be many different foils and the suitable foil(s) will have to be chosen for the context. For example, if task is to predict if the patient will develop Alzheimer’s Disease (fact), then foil may be to predict if the patient will develop Parkinson’s Disease instead (a common misdiagnosis). |
| Explanations are **selective** | People rarely expect a complete explanation consisting of all causes, because the cognitive load of a complete explanation is too large [42]. | Penalize explanations involving too many concepts. | iii) Concepts in explanation and iv) maximum number of concepts | The granularity of features may not necessarily be the apt granularity for an explanation. For example, if we consider a brain MRI, then the features may be 3D voxels. XAI algorithm provides explanation in terms of these voxels, but these can be pooled to different brain regions using brain atlas. Also, high number of concepts in the explanation can put a cognitive burden [42]. |
| Explanations must generate **causal** understanding | The most likely explanation is not necessarily the best because causal reasoning is used to identify if an explanation is satisfactory, not the actual nature of causation [43]. | Restrict our metric to local explanations (explanation for particular prediction rather than about general behaviour of the system). Additionally, contrastive explanations have been associated with generating causal understanding [43], and hence evaluating that helps with this requirement too. | This is satisfied by restricting the scope to local explanations and by evaluating how contrastive the explanation is. |  |
| Explanations are **social** | There are two processes in an explanation: (a) cognitive process - abductive inference is used to deduce the explanation for a given case, and (b) social process - the explanation leads to knowledge transfer between the explainer and the explainee. Thus, we need to consider the expectations of the explainee. | Consideration of clinical context requires scoring explanations using domain-derived concepts (for being relevant to clinicians). These scores for concepts are obtained from biomedical Knowledge Graphs (KG). | iii) Concepts in explanation and v) Knowledge Graph (KG) | The KG needs to be chosen such that it consists of nodes corresponding to the *fact*, *foil*, and the concepts chosen for the explanation. Thus, choice of KG, fact, foil and concepts are interdependent: concepts have to be chosen such that not only the features map to it, but the concepts must also be available in the KG selected. Similarly, the fact and foil nodes, along with relevant relations, must be present in the KG to represent the fact and foil statements correctly. |

**Fig. 1.**
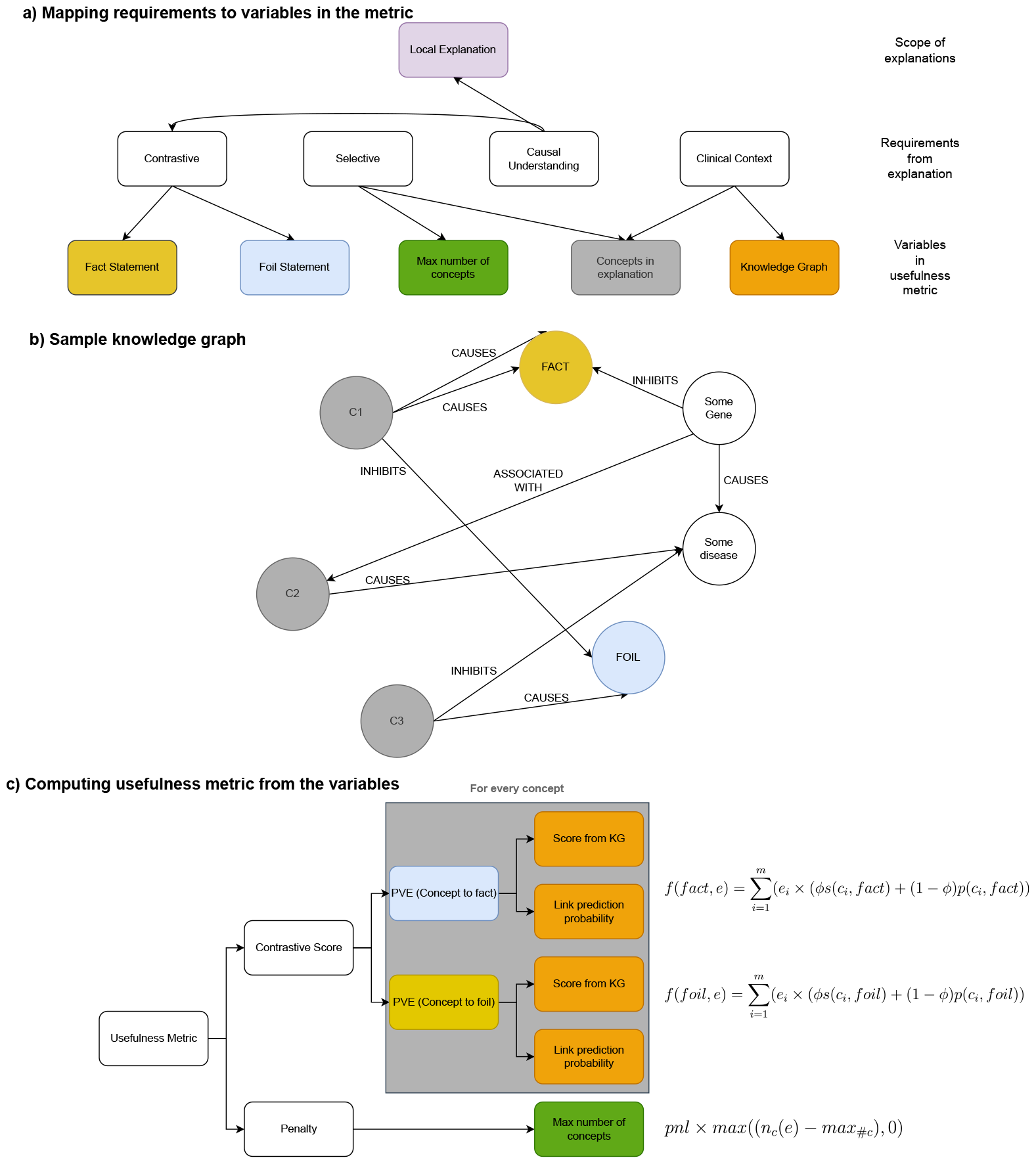
Sample Scoring. The variables of the usefulness metric are shown in (a), a sample knowledge graph (KG) is shown in (b) and the scoring methodology is summarized in (c) and the colors of the variables from (a) are mapped for all derived concepts. The sample KG has the concepts in gray and fact and foil in yellow and blue respectively. Based on the fact and foil statements, the relations of interest can be determined. Consider CAUSES for both fact and foil. In our sample KG (b), edge count scores for the three concepts are [2/4, 0, 1/4]. The link prediction algorithm can provide us an estimate for all the relations, including (C2, CAUSES, fact), even though no edge exists. Both these score are considered using a weighted average to give us the PVE as shown in (c).

Consider an AI predictive system that takes an input *x* (e.g., data or features corresponding to a patient) and has two outputs: the prediction *y* (from an AI model) and the explanation of the prediction represented by *r* (could be from a post-hoc XAI technique or interpretable AI model). We assume the explanation provides an importance score for the features, i.e., *r* is a *k*-dimensional vector where the *i*^th^ dimension, *r*_*i*_, gives the importance score for the *i*^th^ feature (*x*_*i*_) for the particular prediction. Our metric, the *Usefulness* score, is calculated using the following five context-specific variables:

1. Concepts in explanation: Our metric requires the explanation in terms of clinical concepts. If the input features used for the explanation do not correspond to clinical concepts, we sum the feature importance values that map to the same concept to yield concept-level importance values. We represent the set of *m* domain-specific *concepts* by *c* = {*c*_1_, · · ·, *c*_*m*_} and the explanation, at concept granularity, *e* = [*e*_1_, · · ·, *e*_*m*_] by an *m*–dimensional vector, where *e*_*i*_ ∈[ −1, 1], gives the aggregated importance score of concept *c*_*i*_ for the prediction task. To ensure compatibility across explanations from different algorithms, we require *e*_*i*_ to satisfy the condition 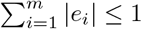.
2. Maximum number of concepts (*max*_#*c*_): We define penalty *p*_#*c*_ = *pnl* × *max*((*n*_*c*_(*e*) − *max*_#*c*_), 0), where *n*_*c*_(*e*) is the number of concepts in the explanation with non-zero importance values *e*_*i*_ and *max*(,) returns the maximum values between its two input variables. Previous studies, e.g., [44], have suggested a maximum of 32 concepts in an explanation for images; but this may vary with context and is taken as an input. The penalty weight *pnl* is the weight by which each additional concept over *max*_#*c*_ is penalized.
3. Fact statement: The fact statement depends on the use-case and the prediction task. E.g., for prediction of development of Alzheimers’ Disease in a patient, the fact statement would be “patient will develop Alzheimers’ Disease”. This fact statement is used to identify the *fact* node in the KG and the relations of interest between the *c*_*i*_ and the *fact*. For example, for our fact statement above, *fact* = Alzheimers’ Disease and relations of interest could be ‘Causes’ or ‘Diagnoses’.
4. Foil statement: Selection of the foil is influenced by the prediction task of the use-case and the fact statement for that use-case. The selection can be done through the following steps.
  a. Consider a fact statement which has k concepts, denoted by (*con*_1_, · · · *con*_*k*_).
  b. For each concept *con*_*i*_ in the fact statement, we derive *candidate* foil statements by considering alternative concepts ¬*con*_*i*_.
  c. If the inherent attributes of a concept *con*_*i*_, can be portrayed as another concept and be derived from the context, then the explanation involving these inherent attributes is called inherence heuristic [45]. E.g., ‘patient’ has an inherent attribute of being in consultation with Neurologist, based on the context. Inherence heuristic can be used as another candidate foil.
  d. The foil statement must be chosen from these candidate foils based on the use-case and would be used to identify the *foil* node in the KG and the relations of interest (between *c*_*i*_ and *foil*) in the KG. Thus, another constraint during foil selection is that the *foil* node must be present in the KG.
5. Knowledge Graph: We use literature-derived KG to calculate the support for a relation between concept *c*_*i*_ and *fact/foil*. To achieve this, we define the following:
  - Concept Score: For a given task *t* (may be *fact* or *foil*), a score indicative of confidence or support for the concept can be calculated from the KG; represented as *s*(*c*_*i*_, *t*). However, not all pairs of concept and task may be connected in the KG, as illustrated in Figure 1(b) and thus *s*(*c*_*i*_, *t*) may be limited by the literature coverage captured in the KG. To address this, we employ link prediction algorithms, e.g., RotatE [46], to provide probabilistic estimates about the association between the concepts and the task, represented as *p*(*c*_*i*_, *t*), even for disconnected pairs in the KG. We use a weighted combination of these two scores for each task: [*ϕs*(*c*_*i*_, *t*) + (1 − *ϕ*)*p*(*c*_*i*_, *t*)], where the weight *ϕ* determines our relative importance to these two scores. The concept score is an estimate of literature support for the concept towards fact in contrast to the foil based on the KG, and is defined as ConceptScore(*c*_*i*_) = [*ϕs*(*c*_*i*_, *fact*) + (1 − *ϕ*)*p*(*c*_*i*_, *fact*)] − [*ϕs*(*c*_*i*_, *foil*) + (1− *ϕ*)*p*(*c*_*i*_, *foil*)]. For example, in SemmedDB KG [47], an edge exists between two nodes (representing clinical concepts) for every pubmed article that connects the said two nodes: more number of edges between two nodes implies more studies containing the relation. So, we can use edge count to calculate *s*(*c*_*i*_, *t*). To determine *ϕ*, we extract the *induced* subgraph, containing the set of nodes of interest (i.e., the concepts, fact and foil nodes) with the edges between these nodes belonging to the set of edge types of interest (i.e., relations of interest from fact and foil statement). The ratio between the degree of fact and foil within the subgraph, in comparison to the entire graph, serves as a measure of how extensively the relations of interest betwen these concepts have been studied. If either the fact or foil is more well-studied for the concepts and relations of interest, then the edge counts-based scores (*s*(*c*_*i*_, *t*)) will be skewed towards well-studied one, making the score less reliable. In that case, we want the *ϕ* to be smaller to give more importance to *p*(*c*_*i*_, *t*). In our case, where the fact is less studied in comparison to the foils, we determine the ra-tio 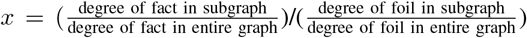, where degree of node is the number of edges incoming and outgoing from the node. Alternatively, if the foil is less studied compared to the fact, we can use 1*/x*. To adjust *ϕ*, we set *ϕ* = (1− *x*)0.5, which means reducing *ϕ* from 0.5 (equal weights) by *x*%. This heuristic can be adapted or altered across KGs and use-cases.
  - Predictive Value of Explanation (PVE): We define a function *f*, called PVE, that uses the explanation (*e*) to weight the scores defined above and determine its predictive value with respect to the fact/foil tasks in KG: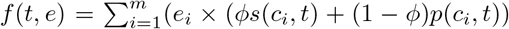. Thus, PVE is higher for explanations which give higher importance values to relations with higher literature support, inferred through the KG.
  - Contrastive Score (CS): As we want the explanation to be more predictive of the *fact* rather than the *foil*, we define a contrastive score 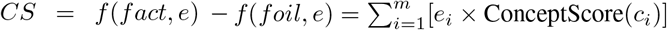.

We define our score, Usefulness = *CS*− *p*_#*c*_. The complete procedure is summarized in Algorithm 1.

#### Algorithm 1 Calculation of usefulness score

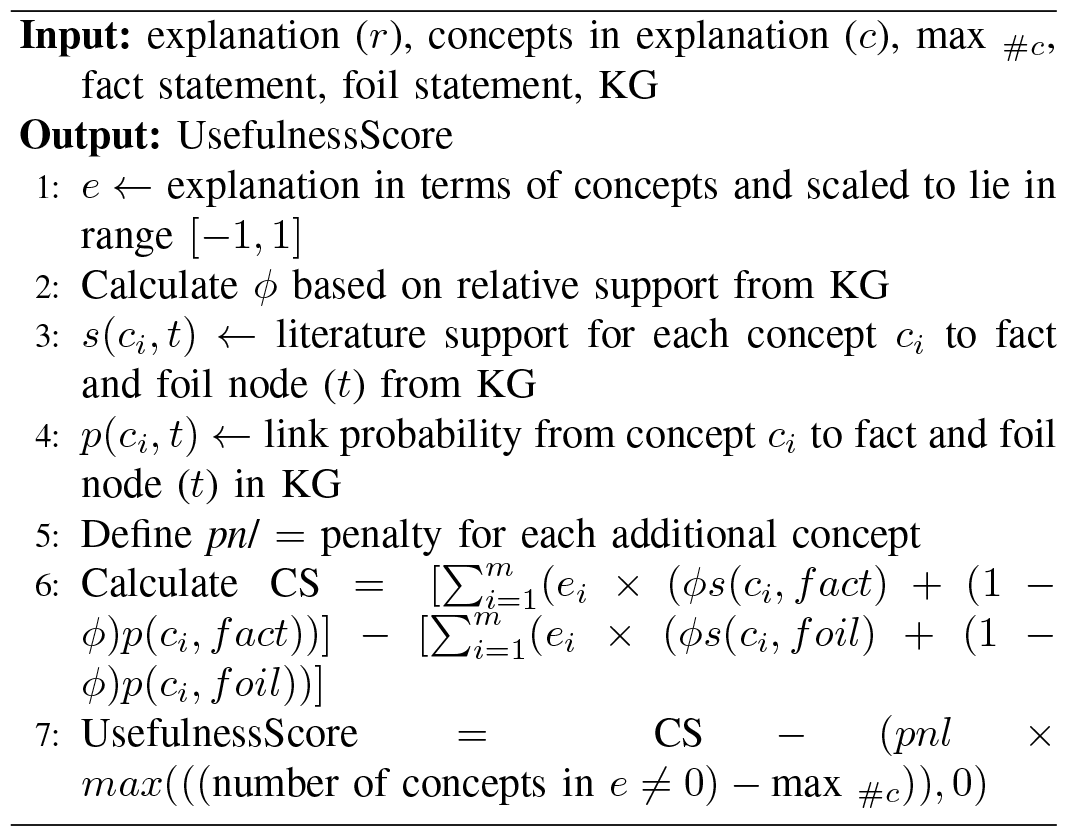

### B. Experiments and Evaluation

#### 1) Context

We consider an early warning system (EWS) for detection of onset of sepsis in ICUs. Sepsis is a life-threatening medical emergency that has high mortality rate; prompt treatment can improve the chances of a successful outcome [48], [49]. The EWS (predictive system) alerts when a patient is predicted to develop sepsis in the next 6 hours (prediction); and may provide information about the patient that led to the prediction (explanation). The application is representative of high-stakes and time-sensitive decision-making which may be supported by predictive algorithms in CDSS [25], [50].

#### 2) Data

We use the data from the PhysioNet Challenge 2019 on the Early Sepsis Prediction [50] and choose the preprocessing strategy and predictive system of a winning team (EASP - Explainable AI Sepsis Predictor) [51]. A total of 37 features are used after all the preprocessing steps. More details can be found in the Appendix I.A.

#### 3) Predictive Systems

For the EWS, we consider 7 different predictive systems. First is an interpretable system (Logistic Regression - LR) that generates predictions with inherent explanations. This produces global explanation, that may also be used to explain prediction for each patient. We compare this with local explanations produced by a post-hoc XAI algorithm - Linear SHAP; called LR+SHAP henceforth. Next, we consider two black-box prediction algorithms: Gradient Boosted Trees (XGB) and a fully connected neural network with 2 hidden layers. The XGB predictions are followed by post-hoc XAI algorithm SHAP (as used by [51]); called XGB+SHAP henceforth. The neural network is followed by 4 post-hoc XAI algorithms – 2 variations of Layer-wise Relevance Propogation (LRP), LRP-0 and LRP-*ϵ* [13], Integrated Gradients (IG) [52] from the innvestigate library [53], and Deep SHAP – called NN+LRP-0, NN+LRP-*ϵ*, NN+IG and NN+SHAP respectively.

#### 4) Experiments

We first qualitatively evaluate our concept score to check its agreement with clinical literature. For this we use the fact – ‘patient is likely to develop sepsis in next 6 hours’ and two different foils – ICU foil ‘patient must be in ICU’ and misdiagnosis foil ‘patient will develop anaphylaxis’ (Table III describes these foils).

**TABLE III.** Choice of variables for case study.

| Variable | Reasoning |  |  |  | Choice |
| --- | --- | --- | --- | --- | --- |
| Concepts in explanation | We use the initial 37 features as concepts because they are various vital signs and laboratory readings. 5 features that do not map to clinical concepts, like the Unit Number, are dropped from concept-level explanation resulting in 33 concepts. There are 168 features in input to the predictive system and consequently in the explanation (see Appendix I.A for details). 131 of these 168 features in the explanation are derived and are mapped back to these 33 concepts for the concept-level explanation. |  |  |  | 33 concepts |
| Maximum number of concepts | [44] identify that most people agree on 32 concepts as a maximum number of concepts in an explanation for images. However, this limit may vary with context and user-groups. As all the 33 concepts in our explanation are important clinical concepts, we define the maximum number of concepts as 33. This implies all explanations generated in this case study will not be penalized. |  |  |  | 33 concepts |
| Fact statement | Based on the objective of the PhysioNet Challenge, we define the fact statement as “patient will develop sepsis in next 6 hours” |  |  |  | ‘patient will develop sepsis in next 6 hours’ |
| Foil statement | Fact Concept | Foil Concept | Candidate Foil statement | Evaluating the candidate foil | ‘the patient must be in ICU’ |
|  | this patient | another patient | This patient will develop sepsis in next 6 hours rather than another patient | The patient foil is not applicable in this case as the KG cannot represent one patient vs the other as two nodes. Also, [24] have also noted that clinicians do not often care about similar patients in this setting. |  |
|  | sepsis | common misdiagnosis like anaphylaxis | The patient will develop some other commonly misdiagnosed disease such as Anaphylaxis | In an ICU, as clinicians have more information and context regarding the patient, [54] recommends identifying the gastrointestinal (GI), genitourinary (GU), or intravenous (IV) source of sepsis. Thus, the diagnosis approach in the ICU is less focused on misdiagnosis. |  |
|  | in next 6 hours | after 6 hours | The patient will develop sepsis after 6 hours | [50] argue that for sepsis prediction, the earlier the diagnosis, better it would be. So, if the system can predict for onset of sepsis more than 6 hours before (time foil), it may not be undesirable, implying explanations in contrast to this foil may not be desirable. |  |
|  | Inherence heuristic | ICU | The patient is in ICU | In the case of ICU patients, if the explanation is in terms of the inherence heuristic, then it would not be valuable as the patient is already critical (hence in ICU) and the doctors also know why [24]. |  |
| Knowledge Graph | SemmedDB [47] is an automatically generated KG using SemRep, a semantic interpreter of biomedical texts, on Pubmed abstracts and citations. It is a heterogeneous graph with different node types and edge types. Node type indicates types of concepts such as Biologically Active Substance, Disease or Syndrome, Cell, etc. Edge types indicate different kinds of associations such as Diagnoses, Causes, etc. The KG is represented as a set of triples (source node, edge, target node). Every node is assigned a specific type to categorize the information it represents. An example triple is: (Leukocytes (type:Cell), Location_of, Necrotizing Vasculitis (type: Disease or Syndrome)). We choose SemmedDB as the KG for our case-study as it contains the concepts and the fact and foil that we have selected above and can provide an estimate of support from clinical literature. To select a link prediction method, we evaluate various graph embedding techniques in the library pykeen [55] on a held-out test set from the KG for the task of predicting a given set of triples. We select the best performing method: RotatE [46] |  |  |  | SemmedDB |

To evaluate our usefulness score, we use the following context-specific variables (i) Concepts: 33 clinical concepts which are the input features – vital signs and laboratory measurements, (ii) *max*_#*c*_ = 33 concept, (iii) Fact statement: ‘patient will develop sepsis in next 6 hours’, (iv) Foil statement: ‘patient is in ICU’, and (v) KG: SemmedDB. Table III shows the reasons for these choices. We compare the 7 predictive systems, mentioned above, using (a) predictive performance, measured using Area Under the Receiver Operating Characteristic (AUROC) on a held-out test set, (b) our usefulness metric and (c) the faithfulness metric [18]. The faithfulness metric incrementally removes the features considered important and verifies its effect on the predictive performance (further details in Appendix I.C.) and has been implemented using AIX360 [56] library. As the explanations generated by LRP-0, LPR-*ϵ* and IG are similar for fully-connected network [57], we expect their usefulness scores to be similar. Additionally, as local explanations have been found to be more useful to clinicians [23], we expect that LR+SHAP will have higher usefulness scores than the global explanation provided by LR.

As another test, we compare the usefulness scores for explanations from a neural network at different stages of training. When the network is not well trained, i.e. upon random initialization, the mapping from features to the predictions are not learnt well which leads to erroneous predictions; the importance values from an XAI algorithm on such a model would also not correlate with features relevant to correct prediction. Thus, a usefulness metric should initially show low scores when the network is not trained and should improve as the network trains.

## IV. Results

### A. Concept scores for sepsis in contrast to misdiagnosis

The concept scores of sepsis in contrast to its commonly misdiagnosed diseases are presented in Table IV, which shows the concepts (row headers) and foils (column headers). These scores also indicate which concepts are helpful in differential diagnosis, e.g., for Pancreatitis foil, Table IV has negative concept score for gender. This implies gender would be more indicative of Pancreatitis than sepsis, and not useful for differential diagnosis of sepsis. We can verify from literature that gender differences in Pancreatitis have been identified [58].

**TABLE IV.** Concept scores for sepsis *contrastive* to commonly misdiagnosed diseases.

| Concept | anaphylaxis | diabetic ketoacidosis | pulmonary embolism | hypovolemia | pancreatitis | vasculitis |
| --- | --- | --- | --- | --- | --- | --- |
| heart rate | 0.0499 | 0.0457 | 0.0303 | 0.0479 | 0.0451 | 0.0421 |
| pulse oximetry | 0.0271 | 0.0111 | 0.0087 | 0.0226 | 0.0061 | 0.0142 |
| temperature | 0.0137 | 0.0119 | 0.0096 | 0.0128 | 0.0087 | 0.0064 |
| systolic blood pressure | 0.0132 | 0.0065 | 0.0085 | 0.0133 | 0.0034 | 0.0093 |
| mean arterial pressure | 0.0061 | 0.0063 | 0.0057 | 0.0066 | 0.0060 | 0.0044 |
| diastolic blood pressure | 0.0067 | 0.0046 | 0.0042 | 0.0059 | 0.0046 | 0.0027 |
| respiration rate | 0.0110 | 0.0140 | 0.0118 | 0.0133 | 0.0119 | 0.0091 |
| end tidal carbon dioxide | 0.0005 | 0.0004 | 0.0003 | 0.0004 | 0.0003 | 0.0002 |
| base excess | 0.0012 | 0.0041 | 0.0036 | 0.0039 | 0.0024 | 0.0033 |
| bicarbonate | 0.0009 | 0.0032 | 0.0030 | 0.0032 | 0.0018 | 0.0024 |
| fraction of inspired oxygen | 0.0003 | 0.0006 | 0.0005 | 0.0006 | 0.0005 | 0.0003 |
| pH | 0.0135 | 0.0041 | 0.0047 | 0.0140 | 0.0034 | 0.0091 |
| partial pressure of carbon dioxide from arterial blood | 0.0024 | 0.0005 | -0.0009 | 0.0029 | -0.0004 | 0.0016 |
| oxygen saturation from arterial blood | 0.0303 | 0.0108 | 0.0111 | 0.0222 | 0.0095 | 0.0161 |
| aspartate transaminase | -0.0034 | 0.0153 | 0.0188 | 0.0214 | -0.0028 | 0.0032 |
| blood urea nitrogen | 0.0116 | -0.0012 | 0.0022 | 0.0117 | -0.0196 | 0.0037 |
| alkaline phosphatase | 0.0072 | 0.0041 | 0.0047 | 0.0058 | -0.0003 | 0.0043 |
| calcium | 0.0151 | -0.0006 | 0.0057 | 0.0177 | -0.0059 | 0.0063 |
| chloride | 0.0127 | 0.0117 | 0.0140 | 0.0226 | 0.0061 | 0.0100 |
| creatinine | 0.0007 | 0.0114 | 0.0144 | 0.0208 | -0.0022 | 0.0064 |
| serum glucose | 0.0001 | 0.0002 | 0.0001 | 0.0001 | 0.0001 | 0.0001 |
| lactic acid | 0.0068 | 0.0062 | 0.0063 | 0.0069 | 0.0063 | 0.0063 |
| magnesium | 0.0257 | 0.0126 | 0.0111 | 0.0211 | 0.0117 | 0.0033 |
| phosphate | 0.0061 | 0.0020 | 0.0022 | 0.0112 | -0.0022 | 0.0032 |
| potassium | 0.0039 | -0.0031 | 0.0047 | 0.0110 | 0.0013 | 0.0043 |
| bilirubin total | 0.0175 | -0.0009 | 0.0050 | 0.0210 | -0.0013 | 0.0012 |
| hematocrit | 0.0220 | 0.0095 | 0.0167 | 0.0298 | -0.0072 | 0.0101 |
| hemoglobin | 0.0147 | 0.0055 | 0.0050 | 0.0136 | -0.0007 | 0.0076 |
| partial thromboplastin time | 0.0078 | 0.0017 | 0.0029 | 0.0080 | 0.0017 | 0.0020 |
| leukocyte | 0.1606 | 0.1325 | 0.1427 | 0.1642 | 0.1282 | 0.1452 |
| platelets | 0.0260 | 0.0039 | 0.0091 | 0.0279 | 0.0046 | 0.0039 |
| age | 0.0640 | 0.0420 | 0.0315 | 0.0600 | 0.0204 | 0.0459 |
| gender | 0.0791 | 0.0681 | 0.0426 | 0.0786 | -0.0171 | 0.0615 |

An important concept for differential diagnosis between sepsis and its mimics (the misdiagnosis) appears to be temperature [59]. Sepsis is commonly observed by an elevated temperature in the patient [54], [59]. Vasculitis typically has elevated temperature as a common complaint, while low-grade temperature might appear in Pancreatitis and patients with Pulmonary Embolism may also have short-lived fever. On the other hand, for Hypovolemia and Diabetic Ketoacidosis, temperature has not been noted in the diagnosis criteria, while for Anaphylaxis, it is noted to be unlikely [59].

As sepsis is also typically accompanied by a high temperature, the clinical concept of temperature is not very indicative of sepsis when considered in contrast to Vasculitis. However, it would be more indicative of sepsis in contrast to Anaphylaxis. The concept score of sepsis in contrast with a disease will indicate how indicative the concept is for sepsis in contrast with the disease. So, we expect the concept score for Vasculitis foil to be lower than for Anaphylaxis foil. In Table IV, we observe that concept score for temperature is lowest for Vasculitis, followed by Pancreatitis, Pulmonary Embolism, Diabetic Ketoacidosis, Hypovolemia and lastly Anaphylaxis. Thus, our concept scores for temperature are in agreement with the literature.

### B. Concept scores for sepsis in contrast to admission to ICU

The concept scores for sepsis in contrast to the ICU foil can be visualized in Fig 2. As per the diagnostic approach for sepsis in ICU [54], hemodynamic parameters, i.e., the cardiac output or blood pressure measurements, are helpful in diagnosing sepsis, but if these are not accompanied by GI, GU or IV disorder, then it is recommended to not assume sepsis; The laboratory concepts considered more indicative of sepsis are leukocytes or platelets. Other concepts in explanations, such as lactic acid, are considered to be useful but not as highly indicative of sepsis diagnosis as abnormality in those concepts could be due to other reasons too. In Fig 2, we observe that the highest score is for leukocytes, while lactic acid has low concept score. Additionally, the factors not mentioned in [54], but indicated in Fig 2 with high concept score are age and gender. Risk factors such as age, gender, and race have been previously identified for sepsis [60]. Thus, the concept scores obtained for sepsis in contrast to the ICU foil are well supported by literature.

**Fig. 2.**
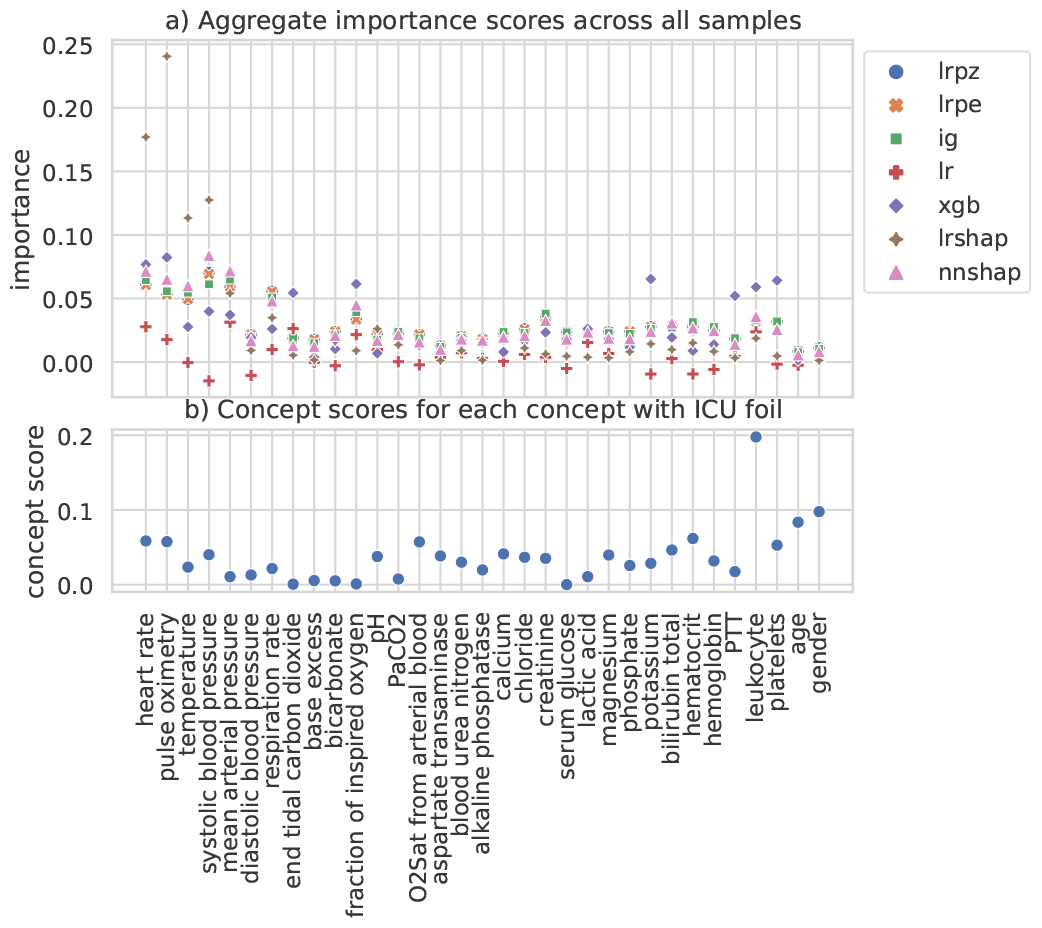
Feature importance of different XAI and the concept scores for sepsis in contrast to ICU foil. a) Mean of the concept importance obtained for each XAI methods (nnshap=NN+SHAP, lrpz=NN+LRP-z, lrpe=NN+LRP-*ϵ*, ig=NN+integrated gradients, lr=logistic regression, lrshap=LR+SHAP, xgb=Gradient Boosted Trees). It is calculated by pooling the features to the mapped concepts. In case of lr, these are simply based on the logits learned during training. b) The concept scores with sepsis as fact and ICU as the foil.

### C. Selection of predictive system

The 7 predictive systems are evaluated. Table V shows our usefulness scores and two other scores – faithfulness and AUROC for the 7 predictive systems we evaluate. Fig 2(a) presents the mean importance score for the explanations generated by each of the predictive systems for all the test samples, and can be used to interpret the usefulness scores in Table V. Though leukocytes is the most indicative concept with highest concept score in Figure 2(b), nearly all the methods give lower importance to this. However, LR has the lowest importance score in most concepts, which indicates why it has a poor usefulness score in Table V. Some concepts such as hematocrit has a high concept score but negative importance score in LR. By the design of our metric, XAI methods that give more importance to concepts with higher concept score will be scored higher in usefulness. Among the predictive systems compared, we observe that XGB+SHAP and LR+SHAP have high mean importance scores for concepts with high concept score. Hence, these have high scores for the usefulness of the explanations generated.

**TABLE V.** Comparison between different CDSS models.

| Method | AUROC | Faithfulness | Usefulness |
| --- | --- | --- | --- |
| **XGB+SHAP** | 0.861257 | -0.116133 | 0.038154 |
| **Logistic Regression** | 0.7564 | 0.13422 | 0.007943 |
| **Logistic Regression+SHAP** | 0.7564 | 0.999916 | 0.044090 |
| **Neural Network+SHAP** | 0.74449 | 0.148981 | 0.036769 |
| **Neural Network+LRP-z** | 0.74449 | 0.034871 | 0.036358 |
| **Neural Network+LRP-ε** | 0.74449 | 0.0320225 | 0.036265 |
| **Neural Network+IG** | 0.74449 | 0.050084 | 0.036226 |

We observe that the XGB+SHAP has the best predictive performance, second-best usefulness score, but the worst faithfulness score. It uses the mean of SHAP values (using TreeSHAP [61]) for 5 Gradient Boosted Trees trained on 5-folds of training data. As noted in [5], TreeSHAP can sometimes produce unintuitive features in the explanation. A mean of these estimates from 5 trees may not be faithful to the actual prediction as indicated by our results. Selecting such a system may potentially bias the clinicians with useful explanations, which are not faithful to the prediction. This may in turn affect the ability of the clinicians to reject incorrect predictions.

As expected, all three XAI techniques - LRP-z, LRP-*ϵ* and IG - have comparable usefulness scores. This shows that our metric does produce similar scores for similar explanations. Additionally, as expected, LR+SHAP has better usefulness scores as compared to LR. This shows that local explanations are given higher scores by our metric as compared to a global explanation, which aligns with prior literature. Thus, although LR+SHAP has lower AUROC compared to XGB+SHAP, overall it has higher faithfulness and usefulness of the explanations provided and could be a good choice considering all three metrics.

### D. Change of usefulness score across training epochs

We plot the usefulness scores at different training epochs of the neural network in NN+LRP-0, NN+LRP-*ϵ*, NN+IG and SHAP in Figure 3. As the networks are trained, their explanations give higher importance scores to features with better predictive signals as compared to explanations from randomly initialized networks. As the networks are trained, we see that the usefulness score of all the 4 post-hoc XAI methods improve significantly over 1000 epochs. Thus, the explanations that give higher importance scores to features with better predictive signals are given higher scores by our metric.

**Fig. 3.**
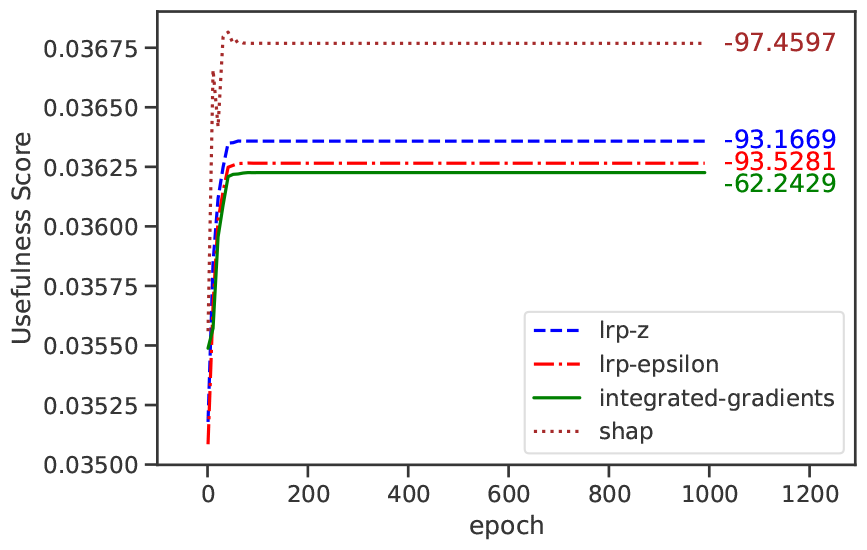
Plot of change in usefulness score along with model training. Text labels at end of the line indicate the t-statistic for the usefulness score between first and last epoch. All four XAI techniques have significant improvement in the usefulness scores at p=0.001.

## V. Discussion

In this paper, we design an evaluation metric to assess the usefulness, in clinical settings, of XAI algorithm explanations, which are in the form of feature importance values. Our experimental results demonstrate the qualities required from such a metric and show that the inference drawn using the metric is in agreement with clinical literature.

Any usefulness metric is highly dependent on and should capture the needs of the user and context. The context-specific variables in our method enables the specification of user and context requirements. As shown by our case-study on sepsis, our metric requires a careful analysis of the context and application to identify the foils and concepts to be used. We expect the use of our metric to help in more user-focused system designs, which in turn may improve the adoption of AI-driven CDSS by clinicians [24]. However, as the CDSS involves use of sensitive patient data, reliability, security and ethical implications must also be considered in practice [1], [62], [63].

Our approach can be used to evaluate any predictive system that provides a prediction and an importance score-based explanation for the features deemed important for the prediction. Thus, any explanation method that can be converted to an importance vector can be used; e.g., decision rules or decision paths may be evaluated using indicator values for the features in the explanation. The usefulness metric can be beneficial, even for an interpretable model such as logistic regression, as our KG-based approach serves as a proxy for how useful the explanations are to clinicians. In some use-cases, multiple foils might be relevant. Our metric can be applied for scoring explanations in contrast to *all* the relevant foils; note that the predictive model need not be trained for all these tasks.

Our methodology is neither dependent on the data used for training nor on the algorithms used for generating the explanation. We use concepts available in biomedical KGs to ensure that the evaluation is in terms of relevant clinical concepts instead of concepts in the data or the algorithms. Thus, an independent source is used to verify the usefulness of the explanations. Our approach can be also adapted to use various model-related elements used for interpretation, such as concept activation vectors [64], as long as these concepts can be mapped to some KG for scoring. Similarly, our methodology can easily be extended to a multi-modal setting with appropriate mapping from features in different modalities to clinical concepts in KGs. For example, the image regions from 3D voxels as well as portions of text notes can be mapped to brain regions (concepts).

A major limitation of our methodology is its dependence on the KG. Literature-derived KGs can be noisy due to errors in the underlying text mining algorithms used to construct the KGs [65] and our method is limited by the correctness of the KG used. Another weakness of our method is the effort required to choose clinically meaningful context-specific inputs (*max*_#*c*_), mapping of concepts to KG and the foils. However, once chosen for a specific setting, the method remains fully automated. Future work can evaluate the impact of these input parameter choices on different problem settings.

As seen in our case study, features in the predictive model may capture temporal characteristics of the data. Our method currently combines such temporal features to the main clinical concept they represent, e.g. the mean heart rate for last 6 hours is mapped to the concept of heart rate. An important area of development would be to extend the metric to explicitly model such temporality.

Our method follows the guidelines for explanations outlined in [39], which are based on commonly accepted criteria in the social sciences. Yet, future research could investigate if specific explanation theories suit particular clinical use cases better. If so, our evaluation method would have to be adapted accordingly. Currently, our method can only evaluate explanations that can be represented by feature importance values, not those obtained interactively from user input. In the future, we aim to expand our method to other types of explanations beyond feature-importance vectors. Furthermore, future studies could examine user interaction with predictive systems across various clinical settings and user groups (e.g., residents, senior consultants, nurses). This research will help refine our evaluation approach by allowing more informed choices for the parameters in our algorithm, such as concept granularity expected and maximum number of concepts.

## VI. Conclusion

We design a new method to evaluate the usefulness of explanations that provide feature importance values from XAI algorithms. To our knowledge, this is the first principled scoring metric to evaluate explanations that is both grounded in the theory of explanations from social science and caters to specific requirements of clinical contexts. Our method presents a novel use of knowledge graphs for choosing clinical concepts at the right granularity for clinicians and for evaluating the value of the clinical concepts used in the explanation. With increasing number of XAI techniques, it becomes important for designers of CDSS to objectively evaluate explanations for use in clinical contexts and our method fulfils this unmet need. Our method is data-type independent and can be used to evaluate explanations from any XAI method that provides feature-based importance values, including post-hoc techniques such as LIME, LRP, SHAP, Saliency map, Integrated Gradients, DeepLIFT or interpretable models like logistic regression or decision trees.

## Data Availability

All data produced in the present study are available upon reasonable request to the authors

## Appendix

### A. Data Preprocessing and Model Training

The PhysioNet data has 40 input features for total population of 40,336 patients. This population is divided into septic and non-septic patients for generating the held-out test data-set. Out of both the population, 15% of the patients are randomly chosen for the held-out test-set on which all evaluations are run.

Out of 40 features for each patient, 3 are dropped due to high number of missing values - direct bilirubin, troponin I, and fibrinogen. For each patient stay, the number of samples equal to the number of hours of stay of the patient are generated by extracting features based on the temporal features of the patient. From the 37 remaining features for each patient sample, 131 features are derived: 62 capture the time interval between measurements, 31 capture difference between previous and current measurement, and 30 capture statistical aggregates of 6-hour sliding windows. The missingness features are extracted as follows: i) informative missingness features reflecting measurement frequency or time interval of raw variables, and ii) differential features, defined by calculating the difference between the current record and the previous measurement value. The remaining missing values are forward filled. This is followed by statistics for 6-hour sliding window for the 5 raw variables: Heart rate, Pulse oximetry, Systolic blood pressure, Mean arterial pressure, Respiration rate. Lastly, 8 features were generated using scoring for value of heart rate (HR), systolic blood pressure (SBP), mean arterial pressure (MAP), respiration rate (Resp), temperature (Temp), creatinine, platelets and total bilirubin according to the scoring systems of NEWS, SOFA and qSOFA. For further details on the procedure and the reasoning, please refer to the original paper.

For the 2 hidden-layer neural network, we use a validation set generated from random 80-20 split on train data for tuning the hyperparameter of the size of first hidden layer. The second layer is fixed to size 10 and the output is 1 neuron.

### B. Concept Mapping

Each concept is mapped to nodes from KG. These concepts were extracted using queries in neo4j. We map nodes to a concept by filtering on the name and label of the node, i.e., all nodes containing the phrase ‘sepsis’ or ‘septic’ such that they were also labeled ‘Disease or Syndrome.’ In some cases, such as in case of chloride, it had to be followed by a manual filtration to remove various chemical compounds. The broader concepts as mentioned in the UMLS were retained and searched while the more specific and narrower concepts were dropped when relevant. For example, for the concept of ‘pulse oximetry’, the blood and venous specific nodes were dropped, but broader concepts of ‘oxygen saturation concept’ or ‘Measurement of oxygen saturation at periphery’ were retained. These decisions were made in accordance with the narrower and broader concepts listed in the UMLS. In most cases, this results in multiple nodes, which are considered together during the calculation of the score.

### C. Faithfulness Metric

The faithfulness metric incrementally removes the features considered important and verifies its effect on the predictive performance. Consider a vector *x* of n dimensions as the input feature vector to the prediction algorithm. Faithfulness is calculated using the Pearson correlation coefficient between the feature importance vector (*f*) and prediction effect vector (*p*) of n dimensions each, where n is the number of features. The vector *p* is defined such that 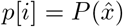, where *P* (.) is function to calculate prediction probability and 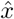 is defined

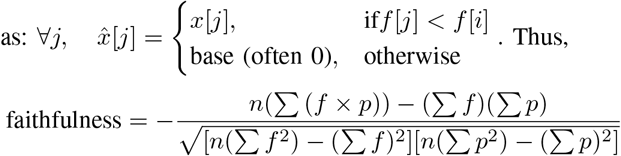

### D. Details on KG specific implementation

In the SemmedDB, a path exists for every document that gives the triple (source node, edge type, destination node). This triple also contains the information of number of times the triple has occured in the document. So, when considering the edge score for a triple, we sum the total number of occurences for all the paths present.

As a concept often maps to multiple nodes, we take the maximum RotatE score from all the nodes. This is because as the KG is formed by automatic extraction, and we also perform automatic extraction, multiple relevant or irrelevant nodes might get picked up, which may be rarely studied. To not bias our scores due to such nodes, we take a max to select the relevant concept which is most studied in the subgraph of our interest.

Additionally, as the score from the RotatE algorithm ranges from [0, −∞), we take an exponential to convert the score to range (0, 1). Similarly, for the edge count, we divide by the maximum count of any node in graph to convert to range (0, 1)

The fact and foil statements are used to select the relations of interest. For our experiments, the relations chosen for fact are: ‘affects’, ‘causes’, ‘diagnoses’, and ‘associated with’. For the foil: ‘affects’, ‘associated with’, ‘occurs in’, ‘location of’, ‘method of’.

### E. Missing features

In practical applications, some features (e.g. location of hospital), may not be present in the KG, restricting their usefulness evaluation. If the features also make sense clinically, then a knowledge graph covering those concepts can be considered; e.g. if brain regions from an atlas are the concepts, then chosen KG must have all these brain regions. In cases like location of hospital, the features may not be mapped to any concept and dropped from the usefulness evaluation.

